# Osteoarthritis across Joint Sites in the Million Veteran Program: Insights from Electronic Health Records and Military Service History

**DOI:** 10.1101/2024.03.01.24303619

**Authors:** Kaleen M Lavin, Joshua S Richman, Merry-Lynn N McDonald, Jasvinder A. Singh

**Author notes:** Correspondence concerning this article should be addressed to: Jasvinder A. Singh, MBBS, MPH, University of Alabama at Birmingham Shelby Building, Room 121, 1825 University Boulevard Birmingham, AL 35233, USA, |. Contributed equally (Merry-Lynn N McDonald, Jasvinder A. Singh).

## Abstract

**Objective:** To characterize the relationship between OA frequency and a host of demographic characteristics, comorbidities, military service history, and physical health variables in a veteran population.

**Methods:** We investigated the Million Veteran Program (MVP) cohort to outline frequency of OA across six joint sites (knee, spine, hip, hand, finger, thumb) in veterans with respect to demographics (age, sex, race/ethnicity, etc.), military service data, and detailed electronic health records profiling OA and other comorbidities. The large veteran contingent provided the unique opportunity to investigate the association of OA with prior service across military branches and war eras.

**Results:** We validated previous reports of sex- and age-dependent differences in OA frequency, and we identified that generalized OA was associated with a higher frequency of sixteen Deyo-Charlson comorbidities. These associations generally persisted within each isolated joint site-specific OA. Depending on military branch, prior military engagement was differentially associated with frequency of OA. Prior Army and Navy service were associated with higher and lower risk, respectively of OA across all joint sites. However, multivariable-adjusted models adjusting for a range of covariates (including age, sex, and ancestry) reversed the apparent protective effect of prior Navy service

**Conclusion:** These findings highlight the breadth of factors associated with OA in the MVP veteran population and suggest that physical status may be a modifiable risk factor for OA. This work may contribute to designing strategies to optimize appropriate detection, intervention, treatment, and even rehabilitation strategies for OA in veterans and the general population.

## INTRODUCTION

Osteoarthritis (OA) is the most prevalent form of arthritis and one of the leading causes of disability in the world^1^. An estimated 14% of the US adult population has OA^2^, and potentially over 250 million are affected worldwide^3^. Prevalence is quickly increasing, especially given the population-level shift towards an older population structure: OA affects about 40% of the world’s population over the age of 70^4^. OA is associated with presence of a range of comorbidities^5^, reduced quality of life, and heightened mortality^6^. The annual economic burden of OA has been estimated to be $89.1 billion^3^, while average medical costs for an individual with OA in the US was $11,542, accounting for both increased insurance costs and cost of OA-related services^2^. The severity and multi-system impact of OA is evident across tissues and includes its characteristic articular cartridge breakdown, synovitis, and inflammatory remodeling of the underlying bone^7^ and overlying skeletal muscle^8^. Furthermore, the cyclical impact of pain and limited mobility on sedentarism and disability contributes to a progressively declining functional state^2^, often persisting even after individuals with OA at operable joint sites (e.g., knee, hip) undergo elective surgery. Risk factors for the development of OA include advancing age, gender, trauma, obesity, genetics, and anatomical differences^1^ but remain incompletely understood.

Current treatment options for OA are limited, with conservative medical treatments providing temporary benefits mostly focused on pain and symptom management or lifestyle interventions that are difficult to sustain^9^. Most individuals with OA eventually elect to undergo joint replacement surgery with progression of the disease^10^ in order to mitigate chronic pain and disability. In the veteran population, a large proportion of individuals continue to experience pain and poor physical function even after surgery and years into rehabilitation^11^. Data even supports that OA-related pain and disability is a central contributor to depression in older veterans^12^.

Large cohort studies such as the Million Veteran Program (MVP) present the opportunity to investigate complex traits such as OA and to examine phenotypic and genetic correlates.^13^ For example, our research team recently leveraged MVP to identify a handful of novel genetic loci robustly associated with OA across genetic ancestry groups, along with validating previously-identified loci and discovering new loci, some of which were uniquely associated to OA in only a given ancestry group^13^. Additionally, the MVP database provides an opportunity to investigate the role of unique lifestyle factors such as prior military service and physical activity. Leveraging this cohort towards a clearer understanding of OA, we investigated multiple factors derived from military service and electronic health records in relation to OA across joint sites in MVP. The purpose of the present investigation was to identify traits related to generalized and site-specific OA in a population of military veterans. It is likely that demographic, genetic, and other measurable factors combine to influence prevalence, progression, and severity of OA.

## METHODS

### Demographics

Participants were curated from the Million Veteran Program (MVP) database. All data were captured between 1990 and 2019. All participants had previously consented to sharing their deidentified data for research. The work described in this manuscript received ethical and study protocol approval from the Veterans Affairs Central Institutional Review Board as well as the University of Alabama at Birmingham (UAB) in accordance with the principles outlined in the Declaration of Helsinki.

OA cases were identified from electronic health record data using International Classification of Diseases (ICD) 9 and 10 codes. Specific ICD codes used for identification of controls, cases with generalized OA, and those with OA across each of six joint sites of interest are provided in previous work by our research team^13^. Inclusion and exclusion criteria were adapted from Zengini, et al.^14^. Briefly, an OA case was defined as an individual whose records had at least one diagnostic code for a generalized OA category, along with at least one diagnostic code for a specific subtype based on the location of the affected joint (e.g., finger, hand, hip, knee, spine, and/or thumb), at least 30 days apart, as has been described.^15^ Veterans presenting with more than one subtype of OA were also included in the data set, as were veterans with prior OA-related joint replacements of the hip or knee. Control participants were identified as those in the dataset that did not have any of the preset case inclusion criteria or any of the control exclusion criteria, such as various non-OA arthropathies. Trauma-based arthropathies were identified by searching for any instance of the string “trauma” in ICD9 and ICD10 codes among those with OA (see **Supplementary Table 9** for complete list of post-traumatic OA ICD codes). Individuals with post-traumatic OA were then filtered from the data set in order to focus on idiopathic OA.

### Military Service

Parameters related to military service were collected, including whether the individual performed any prior service (coded as “active,” “reserves”, “not applicable,” or “missing”), which branch(es) they served in (e.g., Army, Marine Corps, National Guard, etc.), and whether no prior service data were available (importantly, MVP participants may also include veterans’ spouses without any active service history). Calendar years of military service were collected and divided into “eras” based on the primary United States military focus of the associated time period (e.g., WWII, Korean War, Persian Gulf War, modern War on Afghanistan/ Iraq). In order to enable positive responses to multiple options in both of these categories (e.g., served in more than one branch and/or era), as well as to enable an analysis with a higher sensitivity, each of these responses was coded as binary (yes/no) and tested as a separate subcategory.

### Comorbidities and Health Status

The presence of 17 comorbid conditions across body systems were assessed for Veterans in both OA cases and control groups. Conditions included unspecified diseases of perivascular, cerebrovascular, renal, and pulmonary origin, as well as cancer, human immunodeficiency virus / acquired immunodeficiency syndrome HIV/AIDS, diabetes, and others shown in tables below. In order to quantify the collective burden of comorbidities, the commonly-used Deyo-Charlson Comorbidities Index (DCCI) was applied^16^, and Veterans were grouped into categories based on DCCI scores of 0, 1, 2, or ≥3. Veterans were also asked to report the physical demand of their present job and to rate their overall fitness status.

### Statistical Tests

All analysis was performed on the Veterans Affairs Informatics and Computing Infrastructure (VINCI) platform using SQL Studio and R Studio (R version 4.3.1). All numeric data are presented as mean ± standard deviation (SD) and categorial variables as total n and percent relative to the entire group.

#### Univariate Modeling

For numerical values (e.g., age and DCCI), a student’s T-test was performed to compare values between controls and OA cases. For more nuanced examination, age was subdivided into 10-year-span categories. All categorical and binary (yes/no) outcomes were compared between cases and controls using a Chi square goodness of fit analysis of proportions. Each categorical characteristic was also compared individually between controls and site-specific OA (i.e., for the purposes of this investigation, joint sites were not directly compared to one another). This was done in order to establish whether a given joint might be differentially associated with a demographic characteristic in comparison to a control condition. Significant differences were declared at p<0.05, and statistical trends (p<0.10) were acknowledged.

#### Multivariable-Adjusted Modeling

To investigate the independent influence of each variable of interest on the likelihood of generalized and site-specific OA, multivariable-adjusted models were constructed. Variables included in the multivariable-adjusted models were age group, sex, ancestry, body mass index, DCCI score, military branch, and military service type. Additional models were constructed to examine interactions with ancestry and sex with each variable of interest as well sex-stratified and ancestry-stratified models.

## RESULTS

### Generalized OA: Descriptives and Unadjusted Estimates

**Table 1** shows the demographic information for generalized OA cases and controls. Each demographic variable was associated with generalized OA, including female sex, African American, American Indian or Alaska Native ancestry, and non-Hispanic/Latino ethnicity. The rates of previous joint replacement were higher in Veterans with generalized OA than in controls for both hip and knee arthroplasties.

**Table 1:**
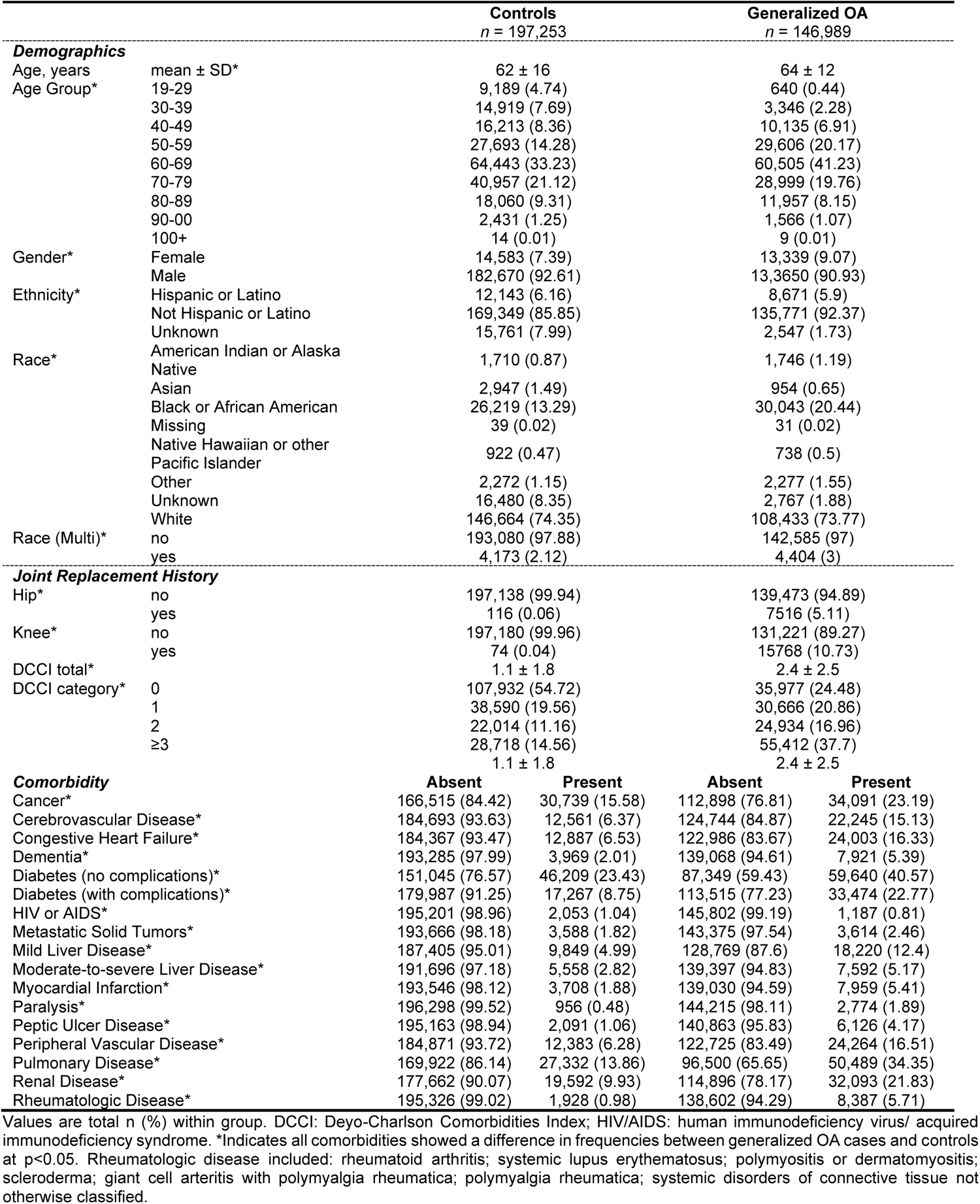
Demographics and Health Status of Generalized OA in the Million Veteran Program (MVP)

Mean Deyo-Charlson comorbidities index (DCCI) was higher (mean ± SD: 2.4 ± 2.5) in veterans with generalized OA than in controls (1.1 ± 1.8) (**Table 1**). Frequency of OA differed across categorical DCCI levels, reflecting this same trend. Specifically, 37.7% of veterans with generalized OA had DCCI category of 3 or higher in contrast with only 14.6% of veteran controls without OA. The presence of 16 of 17 Deyo-Charlson comorbidities was positively associated with generalized OA. The only comorbidity that had a higher frequency among veteran controls than in veterans with OA with generalized was HIV/AIDS.

The frequency of individuals with generalized OA also differed from controls in terms of physical status and physical demand at work (**Table 2**). In general, those with OA reported being in categories of lower fitness and were more likely to report prior active duty or involvement in any military service at all. Veterans with generalized OA were more likely to have served in the Army and National Guard and less likely to have served in the Air Force, Coast Guard, Navy, and multiple branches. A non-significant trend toward this pattern was seen in those who had served in Public Health Service. Veterans with generalized OA were more likely to have served from May 1975 – July 1990 or during the Vietnam era (**Table 2**) but significantly less likely to have served in every other military era except for pre-WWII (November 1941 and earlier), for which no statistical difference was detected. *Generalized OA: Adjusted Estimates*

**Table 2:** Physical Demand, Status, and Military Service in Generalized OA and Control Veterans.

|  | Controls<br>*n* = 197,253 |  | Generalized OA<br>*n* = 146,989 |  |
| --- | --- | --- | --- | --- |
| **Physical Demand at Work\*** |  |  |  |  |
| Very light (mainly sitting) | 39,180 (30.62) |  | 31,720 (32.76) |  |
| Light (mainly walking) | 31,131 (24.33) |  | 19,220 (19.85) |  |
| Medium (lifting, carrying light loads) | 35,494 (27.73) |  | 23,985 (24.77) |  |
| Heavy manual (climbing, carrying heavy loads) | 6,751 (5.28) |  | 5,674 (5.86) |  |
| Missing | 15,420 (12.05) |  | 16,232 (16.76) |  |
| **Current Physical Status\*** |  |  |  |  |
| Very good | 18,414 (14.39) |  | 5,930 (6.12) |  |
| Fairly good | 40,938 (31.99) |  | 20,281 (20.94) |  |
| Satisfactory | 43,295 (33.83) |  | 32,832 (33.91) |  |
| Fairly poor | 20,527 (16.04) |  | 29,245 (30.2) |  |
| Very poor | 3,643 (2.85) |  | 7,461 (7.71) |  |
| Missing | 1,159 (0.91) |  | 1,082 (1.12) |  |
| **Military Service Type\*** |  |  |  |  |
| Active | 120,383 (95.83) |  | 91,526 (96.41) |  |
| Reserves | 2,295 (1.83) |  | 1,303 (1.37) |  |
| N/A | 199 (0.16) |  | 194 (0.2) |  |
| Missing | 2,747 (2.19) |  | 1,914 (2.02) |  |
|  | no | yes | no | yes |
| Missing Service History* | 184,581 (93.58) | 12,672 (6.42) | 145,845 (99.22) | 1,144 (0.78) |
| **Military Branch** |  |  |  |  |
| Air Force* | 101,419 (80.73) | 24,205 (19.27) | 78,381 (82.56) | 16,556 (17.44) |
| Army* | 66,138 (52.65) | 59,486 (47.35) | 46,000 (48.45) | 48,937 (51.55) |
| Coast Guard* | 124,098 (98.79) | 1,526 (1.21) | 93,915 (98.92) | 1,022 (1.08) |
| Marine Corps* | 111,962 (89.12) | 13,662 (10.88) | 83,770 (88.24) | 11,167 (11.76) |
| Merchant Marines | 125,286 (99.73) | 338 (0.27) | 94,665 (99.71) | 272 (0.29) |
| Navy* | 95,751 (76.22) | 29,873 (23.78) | 74,609 (78.59) | 20,328 (21.41) |
| National Guard* | 118,337 (94.2) | 7,287 (5.8) | 89,099 (93.85) | 5,838 (6.15) |
| National Oceanic & Atmospheric Administration | 125,592 (99.97) | 32 (0.03) | 94,908 (99.97) | 29 (0.03) |
| None | 125,622 (100) | 2 (0) | 94,937 (100) | 0 (0) |
| Public Health Service† | 125,460 (99.87) | 164 (0.13) | 94,840 (99.9) | 97 (0.1) |
| Served in Multiple Branches* | 152,288 (77.2) | 44,965 (22.8) | 118,423 (80.57) | 28,566 (19.43) |
| **Military Service Era** |  |  |  |  |
| Sept 2001 or later* | 171,343 (86.86) | 25,910 (13.14) | 136,118 (92.6) | 10,870 (7.4) |
| Aug 1990 - Aug 2001 (incl. Persian Gulf War)* | 152,035 (77.08) | 45,218 (22.92) | 118,991 (80.95) | 27,998 (19.05) |
| May 1975 - Jul 1990* | 155,842 (79.01) | 41,411 (20.99) | 110,772 (75.36) | 36,217 (24.64) |
| Aug 1964 - Apr 1975 (Vietnam era)* | 107,607 (54.55) | 89,646 (45.45) | 68,544 (46.63) | 78,445 (53.37) |
| Feb 1955 - Jul 1964* | 174,480 (88.45) | 22,773 (11.55) | 131,022 (89.14) | 15,967 (10.86) |
| Jul 1950 - Jan 1955 (Korean War)* | 182,718 (92.63) | 14,535 (7.37) | 136,524 (92.88) | 10,465 (7.12) |
| Jan 1947 - Jun 1950* | 195,246 (98.98) | 2,007 (1.02) | 145,658 (99.09) | 1,331 (0.91) |
| Dec 1941 - Dec 1946 (WWII)* | 191,213 (96.94) | 6,040 (3.06) | 142,710 (97.09) | 4,279 (2.91) |
| Nov 1941 or earlier | 197,033 (99.89) | 220 (0.11) | 146,826 (99.89) | 162 (0.11) |

Values are total n (%) within group. OA: Osteoarthritis. \*Indicates $p < 0.05$ , and † indicates $p < 0.10$ toward difference in proportion of Veterans in each category between generalized OA cases and controls via chi square goodness of fit test.

In general, significant associations from the univariate analyses were robust in multivariable-adjusted models adjusting for age group, sex, ancestry, body mass index, DCCI score, military branch, and military service type. Independent of all other factors, frequency of OA was higher in older age groups relative to the youngest group studied. However, risk was modified such that those in age group 50-59, 60-69, and 100+ years had the highest risks of OA (OR = 6.1, 4.8, and 5.3, respectively), but those aged 70-99 years experienced a relatively attenuated higher OA risk (OR ∼3.1). These associations persisted when the data were stratified by ancestry and sex, but no interactions were found. Body mass index (BMI) showed a mild but significant relationship with the risk of OA (OR = 1.05) that persisted for both ancestry- and sex-adjusted and stratified models. Likewise, the heightened OA risk associated with higher comorbidity burden (as indicated by DCCI) was preserved across both sexes and ancestries. None of the unadjusted associations with military branch type were changed when stratified for race or sex (**Supplementary Table 2**).

### Site-Specific OA: Descriptives and Unadjusted Estimates

In the present MVP data set, OA was most and least frequent at the knee and thumb joints, accounting for approximately 22.6% and 1.1% of MVP veterans, respectively (**Table 3**). As **Figure 1** shows, there were often multiple occurrences of OA at joint sites within an individual. This was especially true for hand, finger, and thumb OA. **Figure 1** also shows the large number of individuals that had medical records with generalized OA and no additional data about site-specific subtype. Frequency of OA across all demographic categories (age, sex, race/ethnicity) varied within each site relative to the control group, generally in the direction of the trends seen for generalized OA. The proportion of Veterans who had undergone prior joint replacement history was significantly different within each site vs. control (**Table 3**). While joint sites were not compared to one another, the overwhelming majority of prior joint replacement for operable joints occurred within their respective sites (20% for knee and 30% for hip).

**Figure 1:**
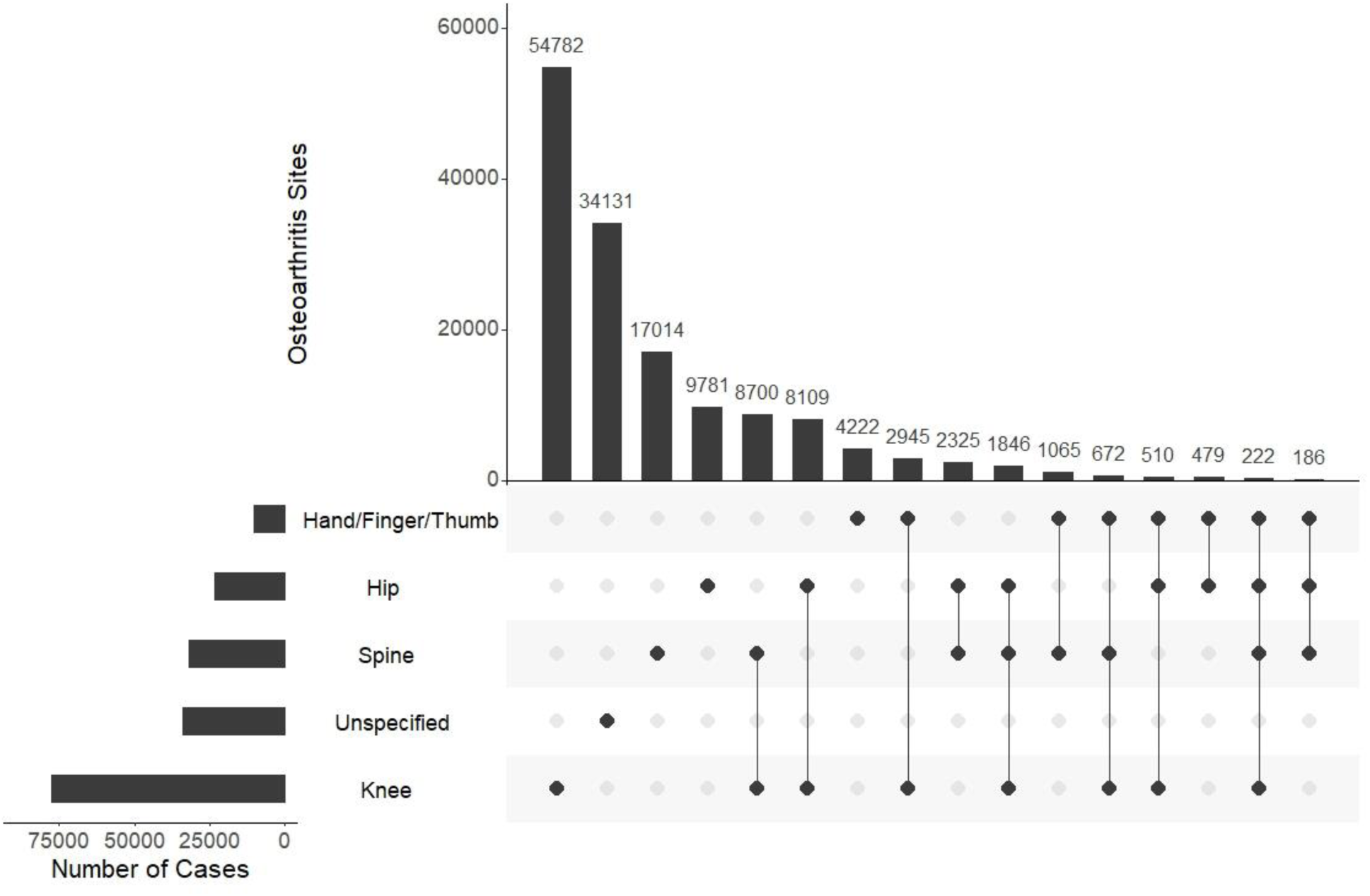
UpsetR plot showing the frequencies of site-specific OA in the Million Veteran Program. Overlap across sites is represented by dot plot on the bottom, where intersecting occurrence of OA at two or more sites is connected by a line. Hand, finger, and thumb OA co-occurred very often and, as such, were collapsed. Knee OA was the most common individual subtype, followed by spine and then hip. Electronic medical records showing generalized OA without a specific site designation are considered “unspecified” and constituted the second-largest group. Combinations of co-occurring site OA are shown by other bars to the right. Only the top 16 intersections are shown for space accommodations.

**Table 3:** Demographics in Site-Specific Osteoarthritis in 344,242 Million Veteran Program Veterans.

|  |  | **Knee OA**<br>*n* = 77,786<br>(22.6%) | **Spine OA**<br>*n* = 32,030<br>(9.3%) | **Hip OA**<br>*n* = 23,458<br>(6.8%) | **Hand OA**<br>*n* = 10,270<br>(3.0%) | **Finger OA**<br>*n* = 8,016<br>(2.3%) | **Thumb OA**<br>*n* = 3,943<br>(1.1%) |
| --- | --- | --- | --- | --- | --- | --- | --- |
| **Demographics** |  |  |  |  |  |  |  |
| **Age, years\*** | mean + SD | 65 ± 11 | 61 ± 12 | 66 ± 11 | 65 ± 10 | 66 ± 10 | 65 ± 9 |
| **Age Group\*** | 19-29 | 171 (0.22) | 297 (0.93) | 26 (0.11) | 10 (0.1) | 7 (0.09) | 3 (0.08) |
|  | 30-39 | 1,191 (1.53) | 1,439 (4.5) | 204 (0.87) | 70 (0.68) | 49 (0.61) | 26 (0.66) |
|  | 40-49 | 4,892 (6.3) | 3,363 (10.52) | 974 (4.16) | 444 (4.33) | 345 (4.31) | 165 (4.19) |
|  | 50-59 | 15,711 (20.23) | 7,733 (24.19) | 4,299 (18.35) | 2,022 (19.71) | 1,543 (19.27) | 843 (21.41) |
|  | 60-69 | 32,665 (42.05) | 12,449 (38.94) | 9,869 (42.12) | 4,714 (45.95) | 3,638 (45.42) | 1,885 (47.88) |
|  | 70-79 | 15,720 (20.24) | 5,025 (15.72) | 5,281 (22.54) | 2,213 (21.57) | 1,766 (22.05) | 806 (20.47) |
|  | 80-89 | 6,495 (8.36) | 1,505 (4.71) | 2,418 (10.32) | 700 (6.82) | 583 (7.28) | 193 (4.9) |
|  | 90-00 | 828 (1.07) | 156 (0.49) | 359 (1.53) | 87 (0.85) | 78 (0.97) | 16 (0.41) |
|  | 100+ | 3 (0) | 2 (0.01) | 2 (0.01) | 0 (0) | 0 (0) | 0 (0) |
| **Gender\*** | Female | 7,374 (9.48) | 3,268 (10.2) | 1,845 (7.87) | 1,385 (13.49) | 1,044 (13.02) | 614 (15.57) |
|  | Male | 70,412 (90.52) | 28,762 (89.8) | 21,613 (92.13) | 8,885 (86.51) | 6,972 (86.98) | 3,329 (84.43) |
| **Ethnicity\*** | Hispanic or Latino | 4,791 (6.16) | 2,135 (6.67) | 999 (4.26) | 584 (5.69) | 451 (5.63) | 218 (5.53) |
|  | Not Hispanic or Latino | 71,663 (92.13) | 29,172 (91.08) | 22,139 (94.38) | 9,513 (92.63) | 7,426 (92.64) | 3,665 (92.95) |
|  | Unknown | 1,332 (1.71) | 723 (2.26) | 320 (1.36) | 173 (1.68) | 139 (1.73) | 60 (1.52) |
| **Race\*** | American Indian or Alaska Native | 976 (1.25) | 421 (1.31) | 226 (0.96) | 99 (0.96) | 80 (1) | 39 (0.99) |
|  | Asian | 458 (0.59) | 280 (0.87) | 100 (0.43) | 85 (0.83) | 70 (0.87) | 26 (0.66) |
|  | Black or African American | 17,377 (22.34) | 6,768 (21.13) | 5,106 (21.77) | 1,431 (13.93) | 1,140 (14.22) | 470 (11.92) |
|  | Missing | 23 (0.03) | 6 (0.02) | 3 (0.01) | 2 (0.02) | 2 (0.02) | 0 (0) |
|  | Native Hawaiian / other Pacific Islander | 421 (0.54) | 171 (0.53) | 89 (0.38) | 44 (0.43) | 34 (0.42) | 16 (0.41) |
|  | Other | 1,267 (1.63) | 582 (1.82) | 280 (1.19) | 160 (1.56) | 132 (1.65) | 61 (1.55) |
|  | Unknown | 1,433 (1.84) | 744 (2.32) | 346 (1.47) | 197 (1.92) | 161 (2.01) | 65 (1.65) |
|  | White | 55,831 (71.78) | 23,058 (71.99) | 17,308 (73.78) | 8,252 (80.35) | 6,397 (79.8) | 3,266 (82.83) |
| **Race (Multi)\*** | no | 75,449 (97) | 30,983 (96.73) | 22,798 (97.19) | 9,944 (96.83) | 7,766 (96.88) | 3,816 (96.78) |
|  | yes | 2,337 (3) | 1,047 (3.27) | 660 (2.81) | 326 (3.17) | 250 (3.12) | 127 (3.22) |
| **Joint Replacement History** |  |  |  |  |  |  |  |
| **Hip\*** | no | 74,560 (95.85) | 30,976 (96.71) | 16,613 (70.82) | 9,954 (96.92) | 7,780 (97.06) | 3,819 (96.86) |
|  | yes | 3,226 (4.15) | 1,054 (3.29) | 6,845 (29.18) | 316 (3.08) | 236 (2.94) | 124 (3.14) |
| **Knee\*** | no | 62,408 (80.23) | 29,905 (93.37) | 21,120 (90.03) | 9,381 (91.34) | 7,361 (91.83) | 3,544 (89.88) |
|  | yes | 15,378 (19.77) | 2,125 (6.63) | 2,338 (9.97) | 889 (8.66) | 655 (8.17) | 399 (10.12) |

As with generalized OA, mean DCCI was significantly higher for each site-specific OA (2.5–3.0-fold higher proportion than controls), and the distribution across DCCI severity subcategories also differed (**Supplementary Table 1**). Of the 17 comorbidities assessed, most reflected the patterns seen in generalized OA. Veterans with OA at all sites other than hip (i.e., knee, spine, head, finger, and spine) were less likely to have HIV/AIDS than controls (values in **Table 1**). The frequency of metastatic solid tumors and OA was higher among veterans with knee and hip OA but no other sites. For each site-specific type of OA, the proportion of individuals differed across physical demand at work categories vs. controls (**Supplementary Table 1**), such that load-bearing OA sites (e.g., knee, spine, and hip) had a higher proportion of individuals in the “very light” work demand category than controls. Veterans with OA at any site generally reported lower physical fitness than controls.

For knee, hip, hand, and finger OA, generally fewer veterans had been in reserves and more had an active service history than controls. In comparison to controls, rates of prior Army service were higher across all site OA subtypes, and rates of prior Marine Corps service were higher for load-bearing joints (**Table 4**). Rates of prior Navy service, as well as service across the category indicating “multiple military branches,” were lower for all OA subtypes vs. control. Knee OA frequency only was lower in Veterans who had served in the Coast Guard, and there was a lower proportion of Veterans with prior Air Force service for all sites other than thumb.

**Table 4:**
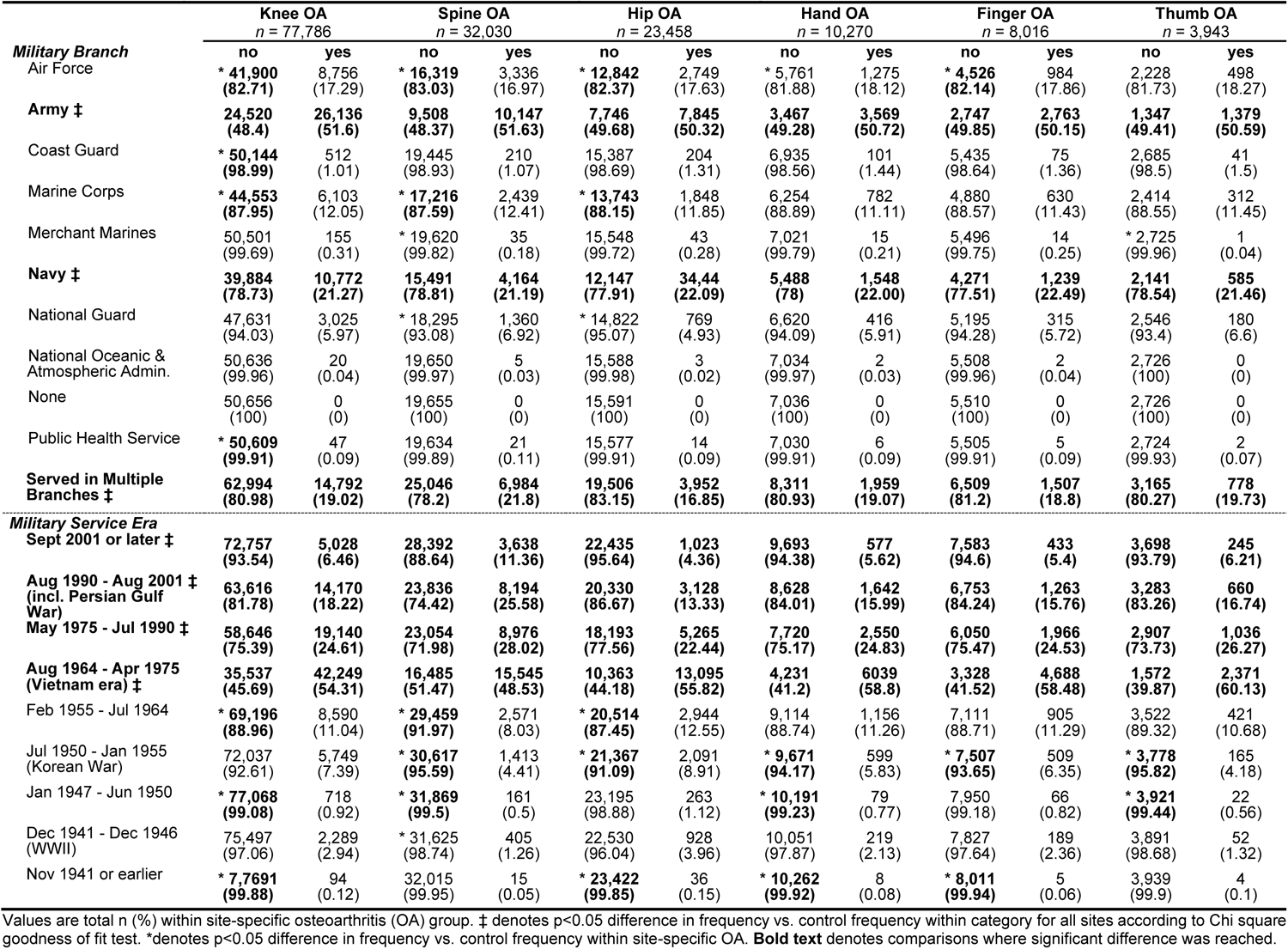
Prior Military Service in Site-Specific Osteoarthritis among Million Veteran Program Veterans.

Across all joint sites, veterans with OA were less likely to have served in a military era after 1990 (**Table 4**). Veterans of Vietnam and the following era tended to have higher frequency of OA at all sites. Other eras presented more variable patterns in site-specific OA (see **Table 4** for details). Briefly, veterans with spine OA were more likely to have served in the Persian Gulf War (25.6%) and World War II (1.3%). Veterans with hip OA were more likely to have served in the Korean War (8.9%) and the following era, whereas other types of site-specific OA tended to be less frequent for those eras. Knee, spine, and hand OA frequencies were lower in veterans who served in the era immediately after WWII. For those with service histories in November 1941 or prior, OA frequency was higher than controls for knee and hip but lower for hand and finger OA.

### Site-Specific OA: Adjusted Estimates

The association between comorbidity burden and risk of OA at all joint sites was upheld in multivariable-adjusted models (**Table 5**). Prior Army service independent of all other factors was associated with odds ratios of 1.2 (knee), 1.1 (hip), 1.3 (hand), and 1.2 (finger) OA. Prior Marine Corps service was also independently associated with odds ratios of 1.5 (spine), 1.4 (knee), and 1.3 (hip). However, the multivariable-adjusted model reversed the apparent protective effect of prior Navy service for spine, hand, and finger OA (OR ∼ 1.1) and eliminated its significance altogether for other joint sites. No significance was found for knee OA frequency in Coast Guard veterans after adjustments in the multivariable models. Additionally, negative associations with prior Air Force service did not persist in the multivariable-adjusted analysis. In fact, spine and hand OA reached significance in the opposite direction (OR ∼1.1). See **Supplementary Tables 2-8** for detailed results of multivariable adjusted models.

**Table 5:**
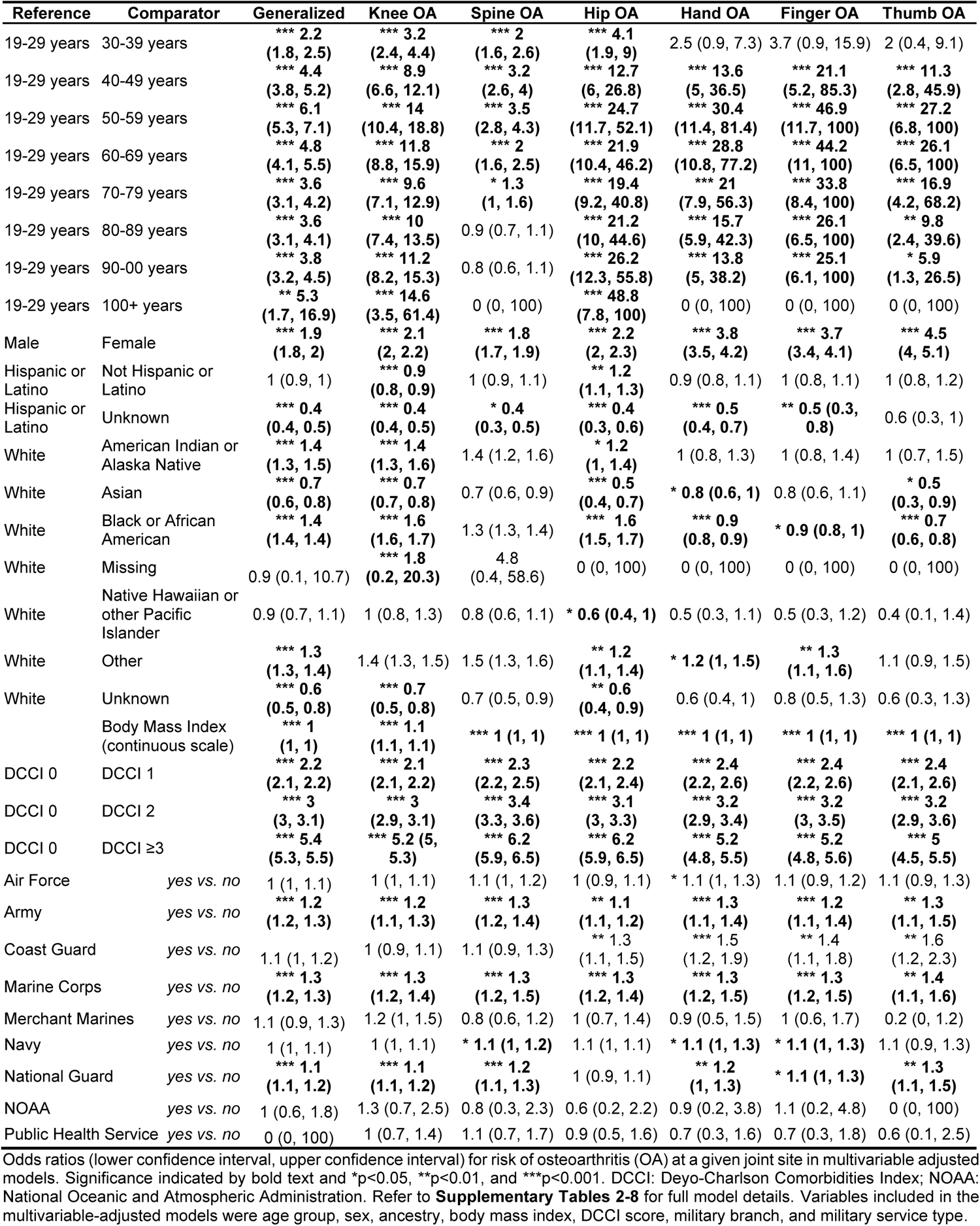
Odds Ratios for Osteoarthritis at Multiple Joint Sites in Million Veteran Program Veterans as Determined by Multivariable Adjusted Models.

## DISCUSSION

OA is a growing public health concern^17^, exacerbated by a rapidly aging population, many of whom have completed military service and may have faced unique environmental exposures, physical challenges, and other triggers.^12^ In the current research, we studied the frequency of OA across joint sites within different demographics and factors in veterans in the MVP. Our findings support the association of sex and ancestry with OA in the veteran population: specifically, female and Black or African American and American Indian or Alaska Native veterans in MVP were more likely to have OA, in keeping with prior reports.^18^ Furthermore, OA is generally accompanied by higher comorbidity burden across multiple physiological systems.

The MVP cohort provides the unique and valuable opportunity to report on the relationship between military service and OA frequency in later life.^13^ In this cohort of veterans, we provide evidence that OA may differentially impact certain military branches and eras, with prior service sometimes showing a protective effect and others a deleterious effect. Veterans with generalized OA, as well as knee and spine OA were more likely to have served in the Army, Marine Corps, or National Guard, compared to other branches. The higher relative frequency of OA in Army, Marine, and National Guard might be due to various exposures that make these Veterans more prone to developing OA.^19,20^ The higher rates of knee and spine OA in these groups may be a product of the more intense physical strain on these joints during service and possibly higher rates of back or limb injury. Active-duty service members in the Army and Marines have a higher rate of injuries and physical trauma compared to Navy and Air Force^21^, which may partially contribute to development of OA.

Multivariable-adjusted model results showed that many service types which appeared partially protective against OA in unadjusted models were not independently significant. This highlights the need to establish a more nuanced understanding of contributors to the relationships across demographics such as age, sex, ancestry, and joint health after service. However, caution should be taken when comparing frequencies across military branches or extrapolating to the general population, as our study did not perform this direct comparison.

Likewise, the differential OA frequencies across military eras are interesting and present potentially novel insight. By necessity, this implicates the known influence of age, as older adults would have served in earlier eras than their younger counterparts. This may partially account for the generally lower frequencies of site-specific OA in more recent war eras vs. higher rates in preceding eras such as the Vietnam War (notably, this era showed the highest proportions of overall service, likely due to implementation of the draft lottery). It is interesting to consider how different site-specific OA subtypes manifest by era, as this also implicates variables outside of the scope of typical examination, such as war strategy, weapon use, fatigue design/weight distribution, and training regimen. For example, spine OA is the most prevalent subtype (based on proportion alone) for those that served in 1990 – 2001. Continued research into the relationship between military era and chronic diseases of interest may provide guidance towards establishing practices that protect joint health and quality of veteran life.

Rates of previous arthroplasty specific to the affected joint were higher in OA cases than in controls for both hip and knee arthroplasties, supporting that OA often manifests bilaterally. This may be due to mechanical (overcompensation, overuse) and/or metabolic (e.g., inflammatory burden) influences, all of which warrant continued investigation^8,22^. This also highlights the importance of appropriately personalized and progressive rehabilitation, as the individual’s unique joint replacement history, gait and biomechanical patterns, and other factors likely converge to impact the success of rehabilitation following joint replacement. In support of this, a recent report suggested that poor recovery to the initial arthroplasty procedure was a major determinant in individuals without military background deciding to cancel a replacement of the contralateral joint, even when medically necessary^23^.

We found that Veterans with OA tended to have jobs that demanded lower levels of physical activity and to report lower overall fitness, but this may be a consequence of OA rather than a cause/determinant of OA. Unfortunately, the progressive pain and disability that accompanies OA often leads to a vicious cycle wherein individuals might reduce daily activity as a strategy to mitigate pain, and the molecular consequences of reduced activity include further muscle weakness, deconditioning, and increased joint pain^8^. The rehabilitation field has focused research efforts toward interventions that might influence this decline, including blood-flow restricted exercise, but continued research is highly warranted^24^.

### Future Directions and Limitations

This study leveraged a large sample size in a richly-phenotyped data set ripe for OA investigation. A limitation of the present work is that OA was identified via electronic health records and not via radiographic assessment, which may lead to misclassification bias. We mitigated this bias by using a previously published algorithm by Zengini et al.^14^ and required the presence of at least two codes at least 30 days apart to establish the presence of OA, which has been shown recently to have high positive predictive value and specificity at the cost of sensitivity.^15^

While the unique military service profile of this large cohort focus has implications for ongoing and future military operations as well as veteran health, care should be taken when generalizing findings. First, participation was voluntary, i.e., not all veterans were included. Secondly, this is by design not a population-level epidemiological investigation and should not be extrapolated to the public. A major reason for this is the overall different demographical composition of this subset of veterans, especially regarding race, ethnicity, and sex. For instance, the proportion of females in the present data set is considerably low (<10%), especially considering that OA is generally more prevalent in females. Insight into OA associations may have been obscured in the present data set due to this low number, particularly when cases were subdivided into joint site. As such, continued investigation is necessary to continue to establish the epidemiology of OA in a population more balanced for sex and ancestry.

The finding that prior military service is associated with differential rates of OA is useful, especially as many branches were related to lower OA frequency during life as a veteran. However, it is difficult to disentangle whether this is causative, or whether these veterans may have been drawn to and selected for active service partially because of their physiological resilience, or “grit”^25,26^. Indeed, it is of high interest of military selection assessments to identify veterans who display advantageous physical and psychological traits such as higher grit and lower susceptibility to overload injury during basic training and subsequent deployment for their own health as well as stability and success of the operation^27,28^. Continued examination of the veteran population is likely to provide critical insight into quality of life following service and may inform existing military protocols of areas of concern.

## Conclusions

With a rapidly aging population structure, accompanied by a shift towards less active lifestyle choices, rates of chronic disease such as OA are expected to rise. Rehabilitation of OA remains highly variable and is too often incompletely successful, and identification of cases is based on chronic pain and loss of normal function. It remains paramount to establish better means of identifying, prehabilitating, and rehabilitating OA across veterans across a wide array of demographic characteristics and personal histories. We provide evidence that these factors, along with joint site specificity, need to be considered in greater detail towards the design of a precision-medicine driven approach to managing OA. Valuable future insight from continued examination of genetics and molecular mechanisms of gene expression regulation along with demographic lifestyle factors may lead to clarity in diagnosing and managing OA. Future studies should consider their important interplay in designing strategies to treat, design pre-habilitation, and even rehabilitate OA across a diverse population.

## Data Availability

All data produced in the present study are available upon reasonable request to the authors.

## AUTHOR CONTRIBUTIONS

Conception and design of research: JAS, MLM; Performed experiments: JR; Drafted manuscript: KML; Prepared figures and tables: JR, KML; Provided feedback on first draft of manuscript: JAS, MLM, JR; Approved final version of manuscript: MLM, JAS, JR, KML

## ACKNOWLEDGMENTS

The authors acknowledge the use of VA CMS in this manuscript.

## VA Million Veteran Program: Core Acknowledgement

### MVP Executive Committee

- Co-Chair: J. Michael Gaziano, M.D., M.P.H. VA Boston Healthcare System, 150 S. Huntington Avenue, Boston, MA 02130
- Co-Chair: Sumitra Muralidhar, Ph.D. US Department of Veterans Affairs, 810 Vermont Avenue NW, Washington, DC 20420
- Rachel Ramoni, D.M.D., Sc.D., Chief VA Research and Development Officer US Department of Veterans Affairs, 810 Vermont Avenue NW, Washington, DC 20420
- Jean Beckham, Ph.D. Durham VA Medical Center, 508 Fulton Street, Durham, NC 27705
- Kyong-Mi Chang, M.D. Philadelphia VA Medical Center, 3900 Woodland Avenue, Philadelphia, PA 19104
- Christopher J. O’Donnell, M.D., M.P.H. VA Boston Healthcare System, 150 S. Huntington Avenue, Boston, MA 02130
- Philip S. Tsao, Ph.D. VA Palo Alto Health Care System, 3801 Miranda Avenue, Palo Alto, CA 94304
- James Breeling, M.D., Ex-Officio US Department of Veterans Affairs, 810 Vermont Avenue NW, Washington, DC 20420
- Grant Huang, Ph.D., Ex-Officio US Department of Veterans Affairs, 810 Vermont Avenue NW, Washington, DC 20420
- Juan P. Casas, M.D., Ph.D., Ex-Officio VA Boston Healthcare System, 150 S. Huntington Avenue, Boston, MA 02130

### MVP Program Office

- Sumitra Muralidhar, Ph.D. US Department of Veterans Affairs, 810 Vermont Avenue NW, Washington, DC 20420
- Jennifer Moser, Ph.D. US Department of Veterans Affairs, 810 Vermont Avenue NW, Washington, DC 20420

### MVP Recruitment/Enrollment

• Recruitment/Enrollment Director/Deputy Director, Boston – Stacey B. Whitbourne, Ph.D.; Jessica V. Brewer, M.P.H. VA Boston Healthcare System, 150 S. Huntington Avenue, Boston, MA 02130
• MVP Coordinating Centers

○ Clinical Epidemiology Research Center (CERC), West Haven – Mihaela Aslan, Ph.D. West Haven VA Medical Center, 950 Campbell Avenue, West Haven, CT 06516
○ Cooperative Studies Program Clinical Research Pharmacy Coordinating Center, Albuquerque – Todd Connor, Pharm.D.; Dean P. Argyres, B.S., M.S. New Mexico VA Health Care System, 1501 San Pedro Drive SE, Albuquerque, NM 87108
○ Genomics Coordinating Center, Palo Alto – Philip S. Tsao, Ph.D. VA Palo Alto Health Care System, 3801 Miranda Avenue, Palo Alto, CA 94304
○ MVP Boston Coordinating Center, Boston-J. Michael Gaziano, M.D., M.P.H. VA Boston Healthcare System, 150 S. Huntington Avenue, Boston, MA 02130
○ MVP Information Center, Canandaigua – Brady Stephens, M.S. Canandaigua VA Medical Center, 400 Fort Hill Avenue, Canandaigua, NY 14424
• VA Central Biorepository, Boston – Mary T. Brophy M.D., M.P.H.; Donald E. Humphries, Ph.D.; Luis E. Selva, Ph.D. VA Boston Healthcare System, 150 S. Huntington Avenue, Boston, MA 02130
• MVP Informatics, Boston – Nhan Do, M.D.; Shahpoor (Alex) Shayan, M.S. VA Boston Healthcare System, 150 S. Huntington Avenue, Boston, MA 02130
• MVP Data Operations/Analytics, Boston – Kelly Cho, M.P.H., Ph.D. VA Boston Healthcare System, 150 S. Huntington Avenue, Boston, MA 02130
• Director of Regulatory Affairs – Lori Churby, B.S. VA Palo Alto Health Care System, 3801 Miranda Avenue, Palo Alto, CA 94304

### MVP Science

- Science Operations – Christopher J. O’Donnell, M.D., M.P.H. VA Boston Healthcare System, 150 S. Huntington Avenue, Boston, MA 02130
- Genomics Core – Christopher J. O’Donnell, M.D., M.P.H.; Saiju Pyarajan Ph.D. VA Boston Healthcare System, 150 S. Huntington Avenue, Boston, MA 02130 Philip S. Tsao, Ph.D. VA Palo Alto Health Care System, 3801 Miranda Avenue, Palo Alto, CA 94304
- Data Core – Kelly Cho, M.P.H, Ph.D. VA Boston Healthcare System, 150 S. Huntington Avenue, Boston, MA 02130
- VA Informatics and Computing Infrastructure (VINCI) – Scott L. DuVall, Ph.D. VA Salt Lake City Health Care System, 500 Foothill Drive, Salt Lake City, UT 84148
- Data and Computational Sciences – Saiju Pyarajan, Ph.D. VA Boston Healthcare System, 150 S. Huntington Avenue, Boston, MA 02130
- Statistical Genetics – Elizabeth Hauser, Ph.D. Durham VA Medical Center, 508 Fulton Street, Durham, NC 27705 Yan Sun, Ph.D. Atlanta VA Medical Center, 1670 Clairmont Road, Decatur, GA 30033 Hongyu Zhao, Ph.D. West Haven VA Medical Center, 950 Campbell Avenue, West Haven, CT 06516

### Current MVP Local Site Investigators

- Atlanta VA Medical Center (Peter Wilson, M.D.) 1670 Clairmont Road, Decatur, GA 30033
- Bay Pines VA Healthcare System (Rachel McArdle, Ph.D.) 10,000 Bay Pines Blvd Bay Pines, FL 33744
- Birmingham VA Medical Center (Louis Dellitalia, M.D.) 700 S. 19th Street, Birmingham AL 35233
- Central Western Massachusetts Healthcare System (Kristin Mattocks, Ph.D., M.P.H.) 421 North Main Street, Leeds, MA 01053
- Cincinnati VA Medical Center (John Harley, M.D., Ph.D.) 3200 Vine Street, Cincinnati, OH 45220
- Clement J. Zablocki VA Medical Center (Jeffrey Whittle, M.D., M.P.H.) 5000 West National Avenue, Milwaukee, WI 53295
- VA Northeast Ohio Healthcare System (Frank Jacono, M.D.) 10701 East Boulevard, Cleveland, OH 44106
- Durham VA Medical Center (Jean Beckham, Ph.D.) 508 Fulton Street, Durham, NC 27705
- Edith Nourse Rogers Memorial Veterans Hospital (John Wells., Ph.D.) 200 Springs Road, Bedford, MA 01730
- Edward Hines, Jr. VA Medical Center (Salvador Gutierrez, M.D.) 5000 South 5th Avenue, Hines, IL 60141
- Veterans Health Care System of the Ozarks (Gretchen Gibson, D.D.S., M.P.H.) 1100 North College Avenue, Fayetteville, AR 72703
- Fargo VA Health Care System (Kimberly Hammer, Ph.D.) 2101 N. Elm, Fargo, ND 58102
- VA Health Care Upstate New York (Laurence Kaminsky, Ph.D.) 113 Holland Avenue, Albany, NY 12208
- New Mexico VA Health Care System (Gerardo Villareal, M.D.) 1501 San Pedro Drive, S.E. Albuquerque, NM 87108
- VA Boston Healthcare System (Scott Kinlay, M.B.B.S., Ph.D.) 150 S. Huntington Avenue, Boston, MA 02130
- VA Western New York Healthcare System (Junzhe Xu, M.D.) 3495 Bailey Avenue, Buffalo, NY 14215-1199
- Ralph H. Johnson VA Medical Center (Mark Hamner, M.D.) 109 Bee Street, Mental Health Research, Charleston, SC 29401
- Columbia VA Health Care System (Roy Mathew, M.D.) 6439 Garners Ferry Road, Columbia, SC 29209
- VA North Texas Health Care System (Sujata Bhushan, M.D.) 4500 S. Lancaster Road, Dallas, TX 75216
- Hampton VA Medical Center (Pran Iruvanti, D.O., Ph.D.) 100 Emancipation Drive, Hampton, VA 23667
- Richmond VA Medical Center (Michael Godschalk, M.D.) 1201 Broad Rock Blvd., Richmond, VA 23249
- Iowa City VA Health Care System (Zuhair Ballas, M.D.) 601 Highway 6 West, Iowa City, IA 52246-2208
- Eastern Oklahoma VA Health Care System (Douglas Ivins, M.D.) 1011 Honor Heights Drive, Muskogee, OK 74401
- James A. Haley Veterans’ Hospital (Stephen Mastorides, M.D.) 13000 Bruce B. Downs Blvd, Tampa, FL 33612
- James H. Quillen VA Medical Center (Jonathan Moorman, M.D., Ph.D.) Corner of Lamont & Veterans Way, Mountain Home, TN 37684
- John D. Dingell VA Medical Center (Saib Gappy, M.D.) 4646 John R Street, Detroit, MI 48201
- Louisville VA Medical Center (Jon Klein, M.D., Ph.D.) 800 Zorn Avenue, Louisville, KY 40206
- Manchester VA Medical Center (Nora Ratcliffe, M.D.) 718 Smyth Road, Manchester, NH 03104
- Miami VA Health Care System (Hermes Florez, M.D., Ph.D.) 1201 NW 16th Street, 11 GRC, Miami FL 33125
- Michael E. DeBakey VA Medical Center (Olaoluwa Okusaga, M.D.) 2002 Holcombe Blvd, Houston, TX 77030
- Minneapolis VA Health Care System (Maureen Murdoch, M.D., M.P.H.) One Veterans Drive, Minneapolis, MN 55417
- N. FL/S. GA Veterans Health System (Peruvemba Sriram, M.D.) 1601 SW Archer Road, Gainesville, FL 32608
- Northport VA Medical Center (Shing Shing Yeh, Ph.D., M.D.) 79 Middleville Road, Northport, NY 11768
- Overton Brooks VA Medical Center (Neeraj Tandon, M.D.) 510 East Stoner Ave, Shreveport, LA 71101
- Philadelphia VA Medical Center (Darshana Jhala, M.D.) 3900 Woodland Avenue, Philadelphia, PA 19104
- Phoenix VA Health Care System (Samuel Aguayo, M.D.) 650 E. Indian School Road, Phoenix, AZ 85012
- Portland VA Medical Center (David Cohen, M.D.) 3710 SW U.S. Veterans Hospital Road, Portland, OR 97239
- Providence VA Medical Center (Satish Sharma, M.D.) 830 Chalkstone Avenue, Providence, RI 02908
- Richard Roudebush VA Medical Center (Suthat Liangpunsakul, M.D., M.P.H.) 1481 West 10th Street, Indianapolis, IN 46202
- Salem VA Medical Center (Kris Ann Oursler, M.D.) 1970 Roanoke Blvd, Salem, VA 24153
- San Francisco VA Health Care System (Mary Whooley, M.D.) 4150 Clement Street, San Francisco, CA 94121
- South Texas Veterans Health Care System (Sunil Ahuja, M.D.) 7400 Merton Minter Boulevard, San Antonio, TX 78229
- Southeast Louisiana Veterans Health Care System (Joseph Constans, Ph.D.) 2400 Canal Street, New Orleans, LA 70119
- Southern Arizona VA Health Care System (Paul Meyer, M.D., Ph.D.) 3601 S 6th Avenue, Tucson, AZ 85723
- Sioux Falls VA Health Care System (Jennifer Greco, M.D.) 2501 W 22nd Street, Sioux Falls, SD 57105
- St. Louis VA Health Care System (Michael Rauchman, M.D.) 915 North Grand Blvd, St. Louis, MO 63106
- Syracuse VA Medical Center (Richard Servatius, Ph.D.) 800 Irving Avenue, Syracuse, NY 13210
- VA Eastern Kansas Health Care System (Melinda Gaddy, Ph.D.) 4101 S 4th Street Trafficway, Leavenworth, KS 66048
- VA Greater Los Angeles Health Care System (Agnes Wallbom, M.D., M.S.) 11301 Wilshire Blvd, Los Angeles, CA 90073
- VA Long Beach Healthcare System (Timothy Morgan, M.D.) 5901 East 7th Street Long Beach, CA 90822
- VA Maine Healthcare System (Todd Stapley, D.O.) 1 VA Center, Augusta, ME 04330
- VA New York Harbor Healthcare System (Scott Sherman, M.D., M.P.H.) 423 East 23rd Street, New York, NY 10010
- VA Pacific Islands Health Care System (George Ross, M.D.) 459 Patterson Rd, Honolulu, HI 96819
- VA Palo Alto Health Care System (Philip Tsao, Ph.D.) 3801 Miranda Avenue, Palo Alto, CA 94304-1290
- VA Pittsburgh Health Care System (Patrick Strollo, Jr., M.D.) University Drive, Pittsburgh, PA 15240
- VA Puget Sound Health Care System (Edward Boyko, M.D.) 1660 S. Columbian Way, Seattle, WA 98108-1597
- VA Salt Lake City Health Care System (Laurence Meyer, M.D., Ph.D.) 500 Foothill Drive, Salt Lake City, UT 84148
- VA San Diego Healthcare System (Samir Gupta, M.D., M.S.C.S.) 3350 La Jolla Village Drive, San Diego, CA 92161
- VA Sierra Nevada Health Care System (Mostaqul Huq, Pharm.D., Ph.D.) 975 Kirman Avenue, Reno, NV 89502
- VA Southern Nevada Healthcare System (Joseph Fayad, M.D.) 6900 North Pecos Road, North Las Vegas, NV 89086
- VA Tennessee Valley Healthcare System (Adriana Hung, M.D., M.P.H.) 1310 24th Avenue, South Nashville, TN 37212
- Washington DC VA Medical Center (Jack Lichy, M.D., Ph.D.) 50 Irving St, Washington, D. C. 20422
- W.G. (Bill) Hefner VA Medical Center (Robin Hurley, M.D.) 1601 Brenner Ave, Salisbury, NC 28144
- White River Junction VA Medical Center (Brooks Robey, M.D.) 163 Veterans Drive, White River Junction, VT 05009
- William S. Middleton Memorial Veterans Hospital (Robert Striker, M.D., Ph.D.) 2500 Overlook Terrace, Madison, WI 53705

